# Emphysema Quantification and Severity Classification with 3-Dimensional Averaging Kernel and Airways Removal

**DOI:** 10.1101/2022.10.31.22281562

**Authors:** Jianxiang Zhang, Gunvant R. Chaudhari, Masha Bonderenko, Jae Ho Sohn

**Affiliations:** University of California San Francisco Medical Center; University of California, San Francisco; University of California San Francisco

## Abstract

**Background:** Emphysema is a common pulmonary pathology known to be associated with increased risk of lung cancer and lung biopsy complications. Prevailing quantitation method of calculating voxel-wise percentage of low attenuation area (LAA) of lung tissue from CT scans is prone to noise and error due overcounting of single voxel LAA and incomplete segmentation of airways.

**Purpose:** We aim to develop an accurate algorithm to quantitatively measure emphysema and classify its severity..

**Methods and Materials:** Two chest CT datasets were obtained from two tertiary hospitals as training and external validation datasets. Exclusion criteria included any patients whose emphysema extent was not specified by the accompanying report. The training dataset included 722 patients, and the validation dataset included 1006 patients. Following lung segmentation and airways removal, we applied convolution of the segmented lung with averaging kernels of different sizes in 2D and 3D. Cutoffs between “none,” “mild to moderate,” and “severe” emphysema were determined via weighted logistic regression on the training dataset, and the categorical emphysema extent was obtained for each patient. The main measure for evaluating model performance was area under the curve (AUC) of the receiver operating characteristic (ROC) on the training dataset and accuracy of classification on both the training and the validation dataset. The 1×1×1 kernel, which is equivalent to the traditional LAA score, was used for comparison to other kernels for performance evaluation.

**Results:** The best model used a 3D 3×3×3 kernel for average filtering with airways post processing and achieved a mean AUC of 0.782 and 0.985 for “none”-versus-rest and “severe”-versus-rest classifications respectively. It achieved a 0.676 and 0.757 multiclass classification accuracy on the training and validation dataset respectively.

**Conclusions and Relevance:** We present an automated pipeline that can achieve accurate emphysema quantification and severity classification. We showed that convolving the segmented lung with a 3D 3×3×3 kernel and post-processing to remove airways can reliably quantify emphysema.

## 1. Introduction

Emphysema is a lung disease belonging to a subtype of chronic obstructive pulmonary disease (COPD) and resulting from the destruction of walls of the acinus (1). COPD was the third leading cause of death in 2019 in the United States (2), with its impact amplified due to the COVID-19 pandemic since then. Furthermore, emphysema has been associated with increased risk and worse prognosis for lung cancer (3–5) and with increased risk of pneumothorax following percutaneous CT-guided lung biopsy (6). Hence, quantification of emphysema can be used for more objective assessment of COPD severity, which can then provide guidance on biopsy planning and interventions such as lung reduction surgery (7,8). Moreover, emphysema quantification allows for a precise and consistent benchmark to incorporate emphysema as a predictor for lung cancer risk stratification and other research projects in pulmonary medicine.

One commonly utilized method for emphysema quantification based on CT imaging is to measure the percentage of voxel-wise low attenuation area (LAA). Previous attempts on emphysema quantification have determined the −950 HU threshold to be optimal or at least near-optimal for voxel-wise LAA classification (9), and LAA scores with the −950 threshold were used in pulmonary function related predictions such as airflow limitation (10). Traditional single voxel-wise LAA is a suboptimal metric for emphysema evaluation because it fails to exclude regions of single voxel LAA that can be physiologically present in patients with normal lung physiology and could be accentuated by quantum mottles especially in low dose chest CTs (11).

Hence, we present an updated lung emphysema quantification algorithm that applies a kernel convolution to account for multiple adjacent patches of LAA and additional image processing to accurately remove airways. We then find the optimal cutoff values for emphysema severity categories by correlating the emphysema quantitation with the corresponding radiologist reported classification.

## 2. Materials and Methods

### Patients

This retrospective, single-institution, institutional review board (IRB) approved, and Health Insurance Portability and Accountability Act (HIPAA)-compliant study included two chest CT datasets. The cut-off determination dataset consisted of consecutively collected 801 patients who were in the lung cancer screening program and obtained chest CT from [hospital 1] from 2015-2022, and data validation dataset consisted of 1093 patients who similarly were in the lung cancer screening program and obtained chest CT from [hospital 2] from 2018-2022. The radiologist-classified emphysema extent for each patient was extracted from corresponding radiology reports using regular expression. Radiology reports that did not mention emphysema were regarded as no emphysema (recorded as “none”). Reports with indications of both emphysema and the extent of emphysema allowed classification of emphysema extent into “mild,” “mild to moderate,” “moderate,” “moderate to severe,” and “severe.” The “mild” and “moderate” classes were combined into “mild to moderate.” and “moderate to severe” and “severe” classes were combined into “severe” for three categories of classification in the end. 89 reports in the cutoff determination dataset and 87 reports in the external validation dataset mentioned emphysema in the report but did not specify the severity, which were therefore noted as “not specified,” and were excluded during the cutoff determination process, as shown in Figure 1.

**Figure 1.**
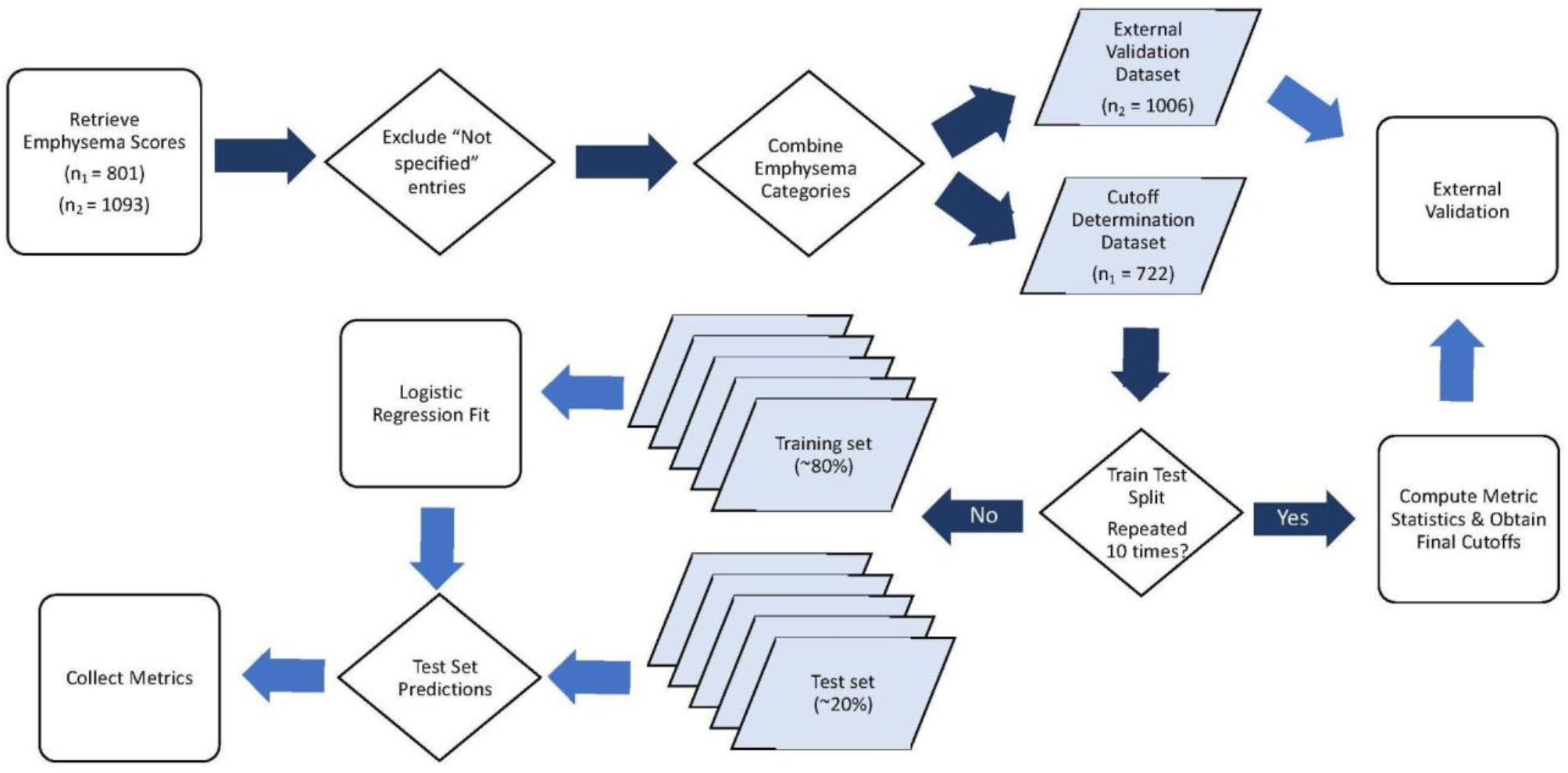
Cutoff determination and validation workflow.

### Preprocessing and Lung Mask Segmentation

For each patient, the full series of axial CT scans was stacked into a 3D array with the first dimension representing the axial direction and following a superior to inferior orientation. The 3D array was first adjusted to a window width of 1500 HU and a window level of −600 HU and resized to 1mm voxels before being passed into an in-house thresholding based segmentation algorithm to retrieve the 3D lung mask. Next, an in-house image processing algorithm was used for segmenting out the lungs while excluding major airways. The axial scans were binarized by a −800 HU threshold, contours of each region were detected, and regions that occupy at least 80% of the entire area enclosed by their contours were flagged as airways and were removed. An illustration of the segmentation algorithm workflow is shown in Figure 2.

**Figure 2.**
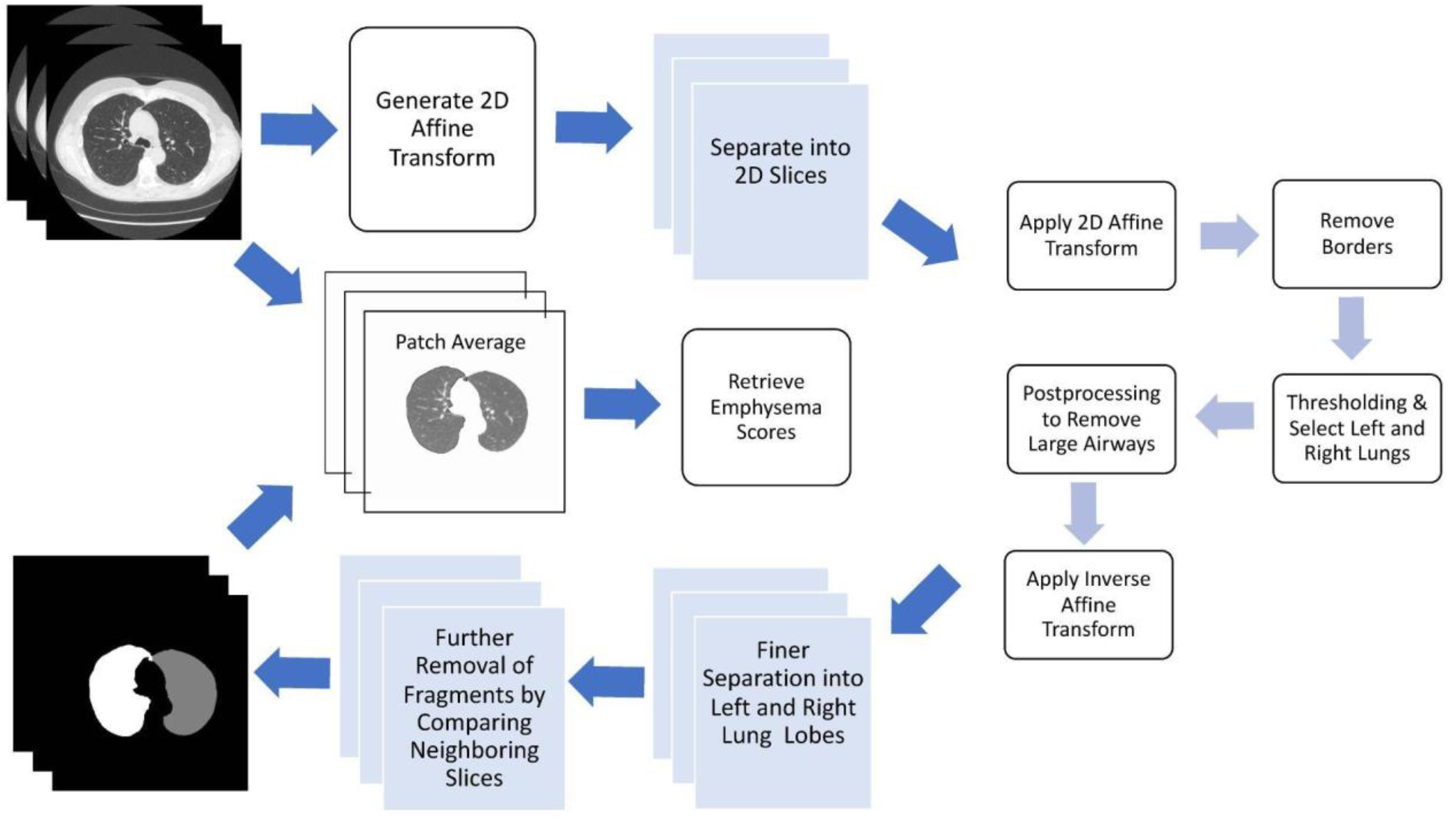
Lung segmentation & emphysema quantification workflow

We tested the accuracy of the lung segmentation and airways removal algorithm on the Lung CT Segmentation Challenge 2017 Dataset, (19–21) (n=60) by comparing the masks outputted by our algorithm and the ground truth masks using Dice score as the metric. Furthermore, 50 randomly selected segmentation masks were reviewed manually by a board certified radiologist (radiologist 1). The source codes for the in-house processing algorithms are made available for review.

### Emphysema Quantification

The windowed image and windowed mask were passed through a function that maped each voxel in the original image to a patch average of a *ps* mm 3D cubic or 2D square kernel, where *ps* is a positive integer, with the original voxel as its center, as shown in the supplementary information section. The emphysema scores are obtained as the fraction of the patch average that is below −950 HU. The patch average obtained from a *ps* =1 kernel in both 2D and 3D cases is equivalent to the original image. Emphysema scores based on 3D kernels of *ps* = 1,3,5 and 2D kernels of *ps* = 3,5,7 were generated for each patient. In practice, any emphysema score larger than 0.100 for all kernels was noted to be severe emphysema.

A board certified radiologist (radiologist 1) also visually evaluated and confirmed the accuracy of the emphysema quantitation masks by visually reviewing emphysema Gaussian-smoothed heat-map overlaid on chest CT images (Figure 3). Only voxels labeled by the algorithm as emphysema-positive were overlaid by a heatmap voxel (shown in the fourth column of Figure 3).

**Figure 3.**
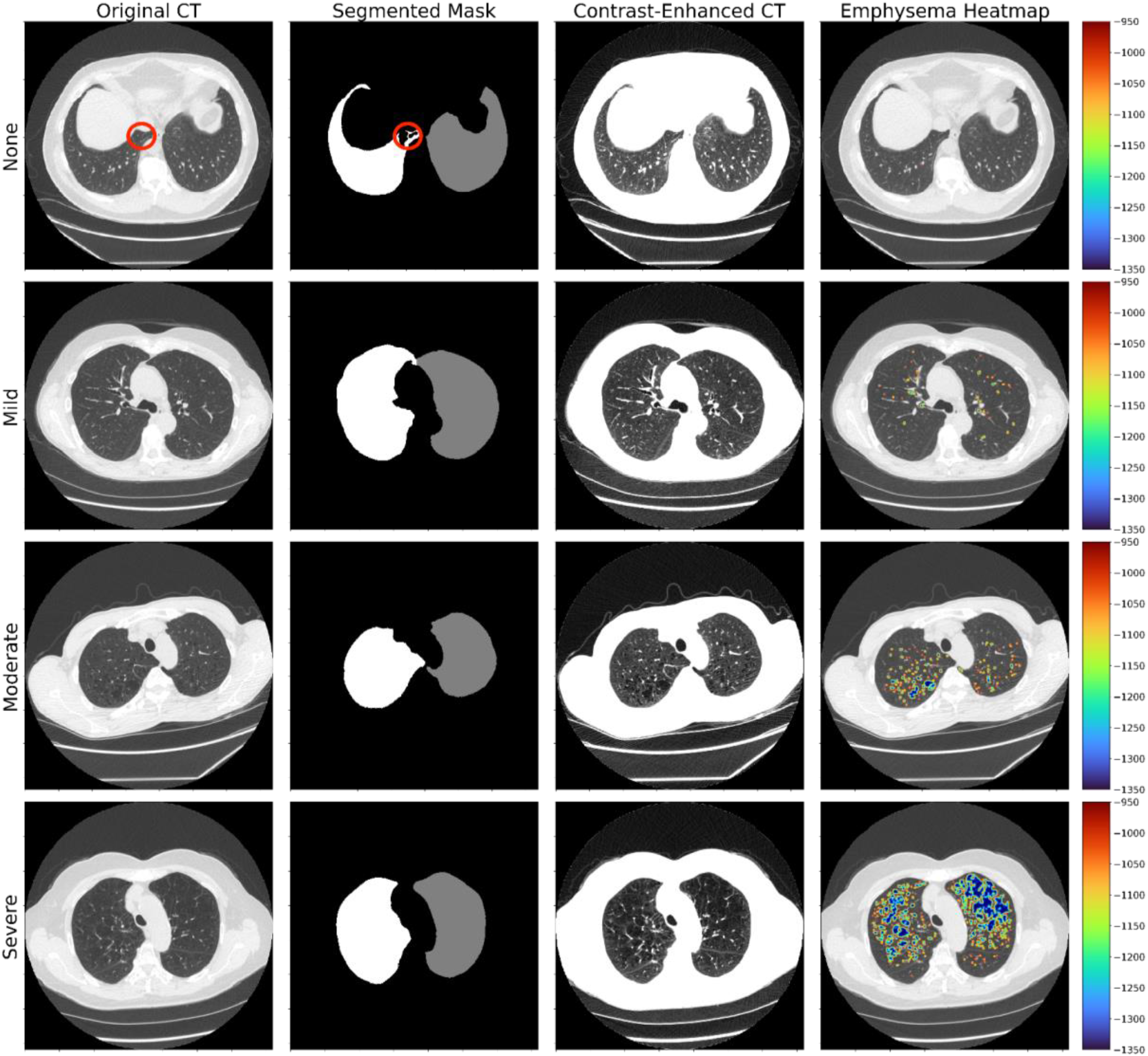
Row 1-4: None, mild, moderate, severe cases. Column 1-4: axial ct scan with window level −600 and window width 1500, lung mask, axial ct scan with window level −750 and window width 600, axial ct scan overlayed with heatmap indicating emphysema locations and severity as classified by 3D kernel with patch size = 3 (i.e. a 3×3×3 kernel). Red circle indicates non-airway regions removed by the postprocessing step of the segmentation algorithm.

### Calibration with Cutoff Determination

A total of 722 chest CTs from [hospital 1] were obtained after excluding “not specified” entries (n=89) to determine the emphysema severity cutoffs. Random split stratified by emphysema category was applied to produce 5 consecutive folds of the dataset, thus generating five pairs of training and test dataset with the test dataset being each fold and the training dataset being the rest 4 folds combined.

For each training set, the outliers for each class, defined by scores that are either greater than upper quartile *(UQ)* + 1.5*interquartile range *(IQR)* or less than lower quartile *(LQ)*-1.5\**IQR*,(12) were removed (13–15). The cutoffs for separating “none” and “mild to moderate” and for separating “mild to moderate” and “severe” were determined by “none”-versus-rest and “severe”-versus-rest binary classifications respectively.

For each classification, a weighted logistic regression model (sklearn 1.1.0 LogisticRegression class) with weights inversely proportional the number of samples in each category was trained on each training set with the emphysema score as the variable and the binarized encoding of the emphysema extent as the ground truth label. An ROC curve was plotted on the model’s prediction on validation set, and the emphysema score cutoff was selected based on maximizing Youden index to optimize the trade-off between sensitivity and specificity (16). The final cutoff was determined, and the emphysema prediction on the test set was made.

Predictions from the five test sets were recombined to give the final predictions for all 722 patient entries. Four metrics — the mean absolute encoded difference between prediction and ground truth labels, the multiclass accuracy, F-score, and kappa score, — were computed. The three categories — “none,” “mild to moderate,” “severe” — were encoded into 0,1,2 respectively to calculate the mean absolute encoded difference.

The above process (from train-test split to metric calculation) were repeated 10 times to obtain AUC distributions (n=50) and metric distributions (n=10). The final cutoffs were determined by averaging across the cutoffs determined from each training set (n=50). The entire cutoff determination workflow is illustrated in Figure 1.

### Validation Dataset

For validation of algorithm performance, 1006 lung cancer screening CTs from [hospital 2] were collected after excluding “not specified” entries (n=87). Emphysema scores were produced using the six kernels, and the corresponding emphysema classifications were made based on the final cutoffs obtained for each kernel from the cutoff determination dataset.

## 3. Results

### Patients

Demographics of the two datasets (after exclusion criteria) are shown in Table 1. 722 participants and 1006 participants were included in the cutoff determination and external validation dataset respectively. Both dataset had an average age in the 60-65 range and had more male participants, with the external validation dataset consisting of more males. Both datasets largely contained patients with no or mild emphysema.

**Table 1.** Demographic statistic of patients after exclusion

|  | **Cutoff**<br>**Determination Dataset** | **External**<br>**Validation Dataset** |
| --- | --- | --- |
| **Number of Participants (Total)** | 722 | 1006 |
| **none** | 472 | 736 |
| **mild** | 164 | 148 |
| **mild to moderate** | 7 | 11 |
| **moderate** | 54 | 76 |
| **moderate to severe** | 5 | 5 |
| **severe** | 20 | 30 |
| **Male %** | 62.2 | 84.6 |
| **Age** | 65.4±6.5 | 63.2±6.6 |

### Preprocessing and Lung Mask Segmentation

Upon testing the accuracy of our lung segmentation algorithm with the Lung CT Segmentation Challenge 2017 Dataset, our algorithm achieved a mean Dice score of 0.930 with a standard deviation of 0.030. The first two columns of Figure 3 present one example of the lung mask outputted by the algorithm in comparison with the original image from each category of none, mild, moderate, severe cases. All 50 out of 50 randomly sampled lung segmentation masks that were reviewed by the board certified radiologist were confirmed to be accurate in identification of the left/right lobe and qualitatively satisfactory in segmentation of the lung parenchyma.

### Emphysema Quantification

As shown in a representative example in Figure 3, the area of heatmap increased drastically with increased emphysema severity according to radiologist labels, indicating that the emphysema quantification algorithm was able to capture emphysema-positive regions and differentiate among lung scans of different emphysema severities. False positive cases were also recorded for error analyses, including pulmonary cysts and honeycombing cases that were erroneously identified as emphysema (Figure S1).

### Calibration with Cutoff Determination

Depending on the kernel and the exact split into the sub-training and validation datasets, the outlier removal decreased the number of patients in the sub-training dataset from n=577-578 to n=477-565. For each cutoff, during the cutoff determination process, 50 AUCs on the validation dataset were obtained for each kernel (10 repeats of 5 train-test split,). Figure 4 compares the distribution of AUCs given by logistic regression for each of the 6 kernels. For 3D kernels, a *ps*=1, which is equivalent to LAA, resulted in lower AUCs for both cutoffs. The three 2D kernels showed similar AUC distributions, with 3×3 and 5×5 performing better for distinguishing “none”-versus-rest emphysema. For both “none”-versus-rest and “severe”-versus-rest classifications, the AUC of *ps*=1 is significantly lower than AUCs of other kernels. The AUC of 3D *ps*=5 is significantly lower than AUCs of other kernels for “none”-versus-res classification. The mean AUCs are also presented in Table 2, with the 3D *ps*=3, 2D *ps*=3, and 2D *ps*=5, 2D *ps*=7 kernels achieving the similar mean AUCs of 0.774-0.794 and 0.985-0.992 for “none”-versus-rest and “none to moderate” versus “severe” classification respectively.

**Table 2.** Quantitative cutoffs (for cubic-root transformed emphysema scores) and metrics for emphysema classification based on different kernels. Abbreviations: LAA (fraction of low attenuation area), ps (patch size of kernel)

|  | **3D ps=1**<br>**(LAA)** | **3D ps=3** | **3D ps=5** | **2D ps=3** | **2D ps=5** | **2D ps=7** |
| --- | --- | --- | --- | --- | --- | --- |
| **Cutoff("none"**<br>**vs "rest")** | 0.280 | 0.159 | 0.082 | 0.131 | 0.094 | 0.057 |
| **Cutoff("severe"**<br>**vs rest)** | 0.377 | 0.261 | 0.158 | 0.254 | 0.162 | 0.135 |
| **AUC ("none"**<br>**vs rest)** | 0.692±0.010 | 0.782±0.013 | 0.712±0.013 | 0.796±0.011 | 0.794±0.009 | 0.774±0.010 |
| **AUC**<br>**("severe"**<br>**versus rest)** | 0.949±0.006 | 0.985±0.002 | 0.987±0.006 | 0.988±0.003 | 0.992±0.002 | 0.991±0.003 |
| **mean**<br>**absolute**<br>**difference** | 0.482±0.008 | 0.365±0.004 | 0.38±0.005 | 0.337±0.005 | 0.355±0.003 | 0.366±0.007 |
| **multiclass**<br>**accuracy** | 0.583±0.005 | 0.667±0.003 | 0.643±0.005 | 0.686±0.004 | 0.675±0.002 | 0.663±0.006 |
| **F score** | 0.577±0.006 | 0.676±0.003 | 0.658±0.007 | 0.702±0.004 | 0.706±0.002 | 0.672±0.009 |
| **kappa score** | 0.196±0.005 | 0.316±0.008 | 0.257±0.009 | 0.333±0.007 | 0.304±0.005 | 0.312±0.008 |

**Figure 4.**
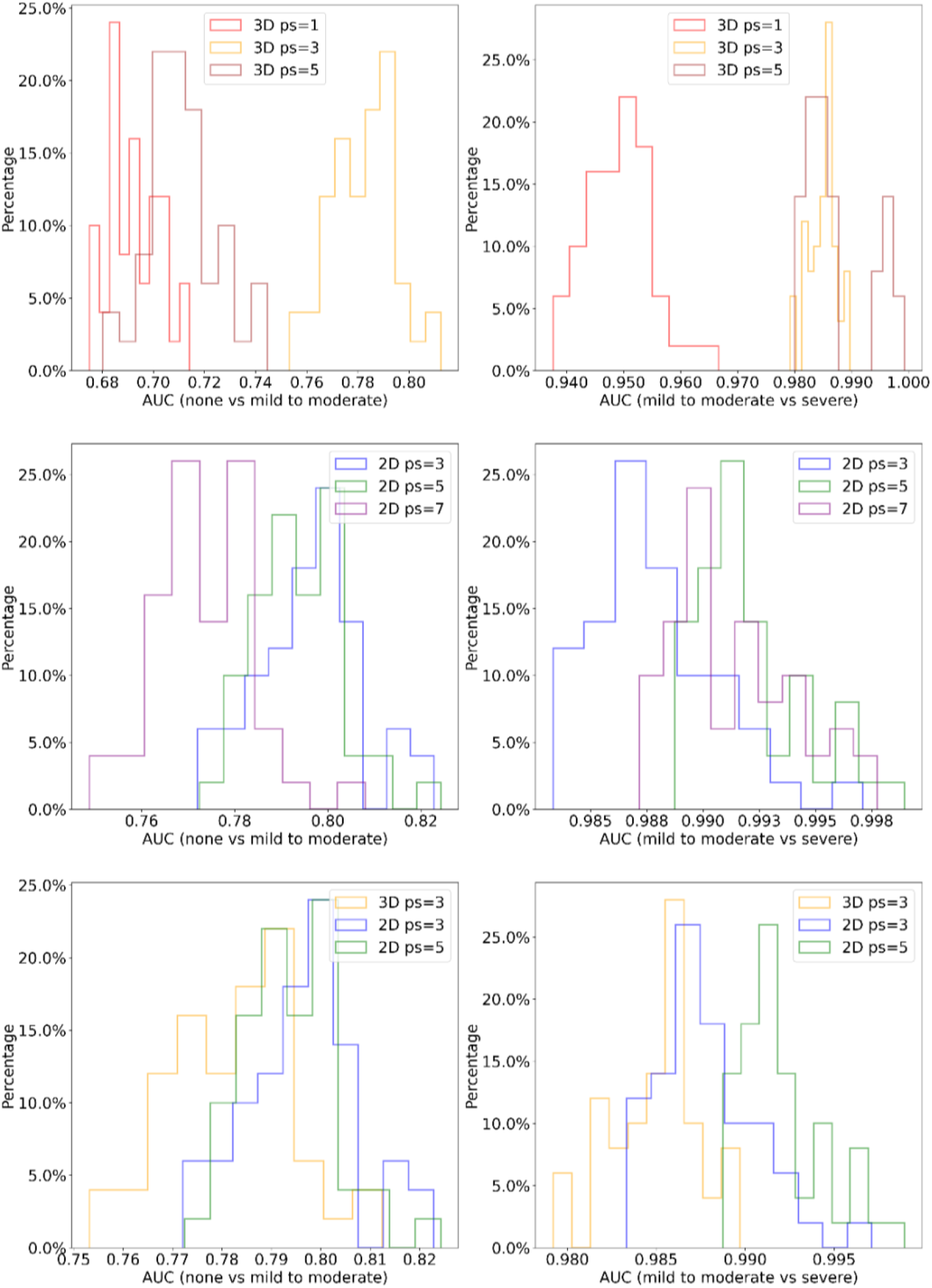
Comparison of AUC distributions for different kernels. LAA score (3D ps=1) and 3D ps=5 kernels had the lowest AUC scores.

For each kernel, boxplots of the emphysema score distributions grouped by radiologist classification in the cutoff determination dataset were plotted in Figure 5, and quantitative metrics were presented in Table 2. As shown in Figure 5A and 5B, the median emphysema scores showed a consistent increase with the increase in emphysema severity for all kernels, with the spread of emphysema score distribution for “none” and “mild” but not “severe” decreases rapidly with kernel size. LAA score had the highest mean absolute difference between model predictions and radiologist classifications (0.482) and the lowest accuracy (0.583). 3D *ps*=5 kernel had the second lowest accuracy (0.643). All other kernels achieved accuracy in range 0.663-0.686, with 2D *ps*=3 and 3D *ps*=3 kernels achieving the highest kappa scores, and 2D *ps*=3 and 2D *ps*=5 kernels achieving the highest F scores. As shown in Figure 5B, all cutoffs demonstrated separations between the three classes.

**Figure 5.**
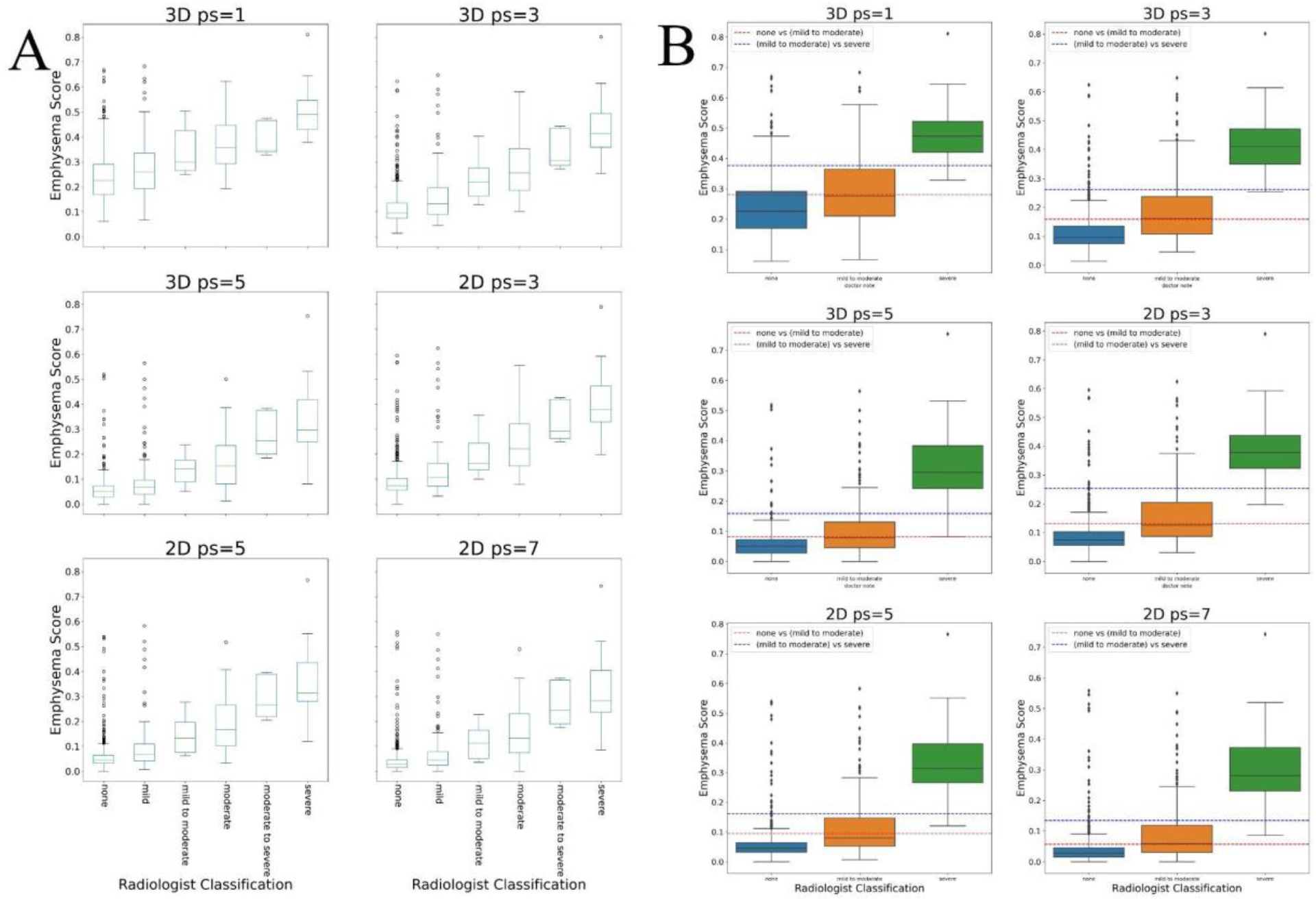
(A) Cubic root transformed emphysema score distributions of the cutoff determination dataset of 6 different kernels. (B) Cubic root transformed emphysema score distributions after regrouping with the final cutoffs.

### External Dataset Validation

Finally, the metrics for ternary (none, mild-moderate, and severe) classification on the external validation dataset are shown in Table 3. Boxplots of emphysema score distributions for each kernel are presented in Figure 6. Comparison between the Figure 5B and Figure 6B demonstrated a disparity in the emphysema score distributions between 3D kernels and 2D kernels. While the two 3D kernels (*ps=3* and *ps=5*) showed slight decrease or remained constant in terms of the spread of score distribution in “none” and “mild to moderate” classes, which was consistent with the LAA score, all three 2D kernels exhibited a larger spread of “none” and “mild to moderate” distributions. All three 2D kernels as well as the 3D *ps*=5 kernel had much higher mean absolute encoded difference (0.354-0.521) and lower accuracy (0.561-0.674) than LAA score (mean difference=0.267, accuracy=0.747). The 3D *ps*=3 is the best kernel with lowest mean difference 0.257, highest accuracy 0.757, and highest kappa score 0.378. Indeed, as shown in Figure 6B, only the 3D *ps*=3 cutoffs showed reasonable separations between the three classes. The confusion matrices for the 3D kernels for the cutoff determination dataset (averaged over 10 repeats) and the external validation dataset are provided in Table S1 and S2.

**Table 3.**
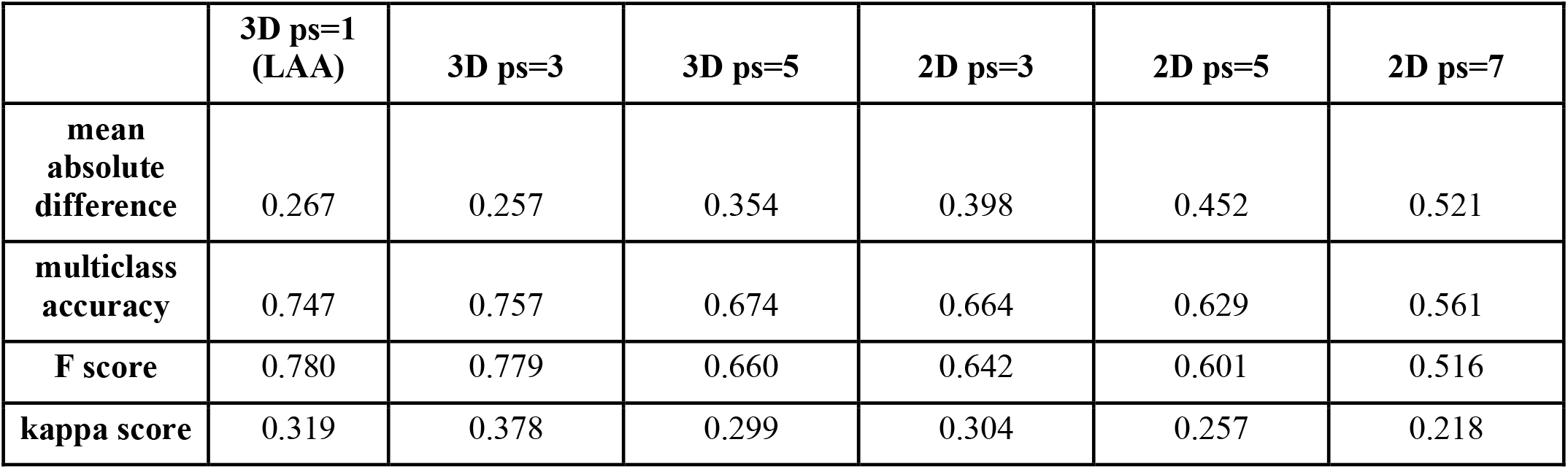
Quantitative metrics for emphysema classification on external validation dataset using final cutoffs obtained from the cutoff determination dataset. Abbreviations: LAA (fraction of low attenuation area), ps (patch size of kernel)

|  | **3D ps=1**<br>**(LAA)** | **3D ps=3** | **3D ps=5** | **2D ps=3** | **2D ps=5** | **2D ps=7** |
| --- | --- | --- | --- | --- | --- | --- |
| **mean**<br>**absolute**<br>**difference** | 0.267 | 0.257 | 0.354 | 0.398 | 0.452 | 0.521 |
| **multiclass**<br>**accuracy** | 0.747 | 0.757 | 0.674 | 0.664 | 0.629 | 0.561 |
| **F score** | 0.780 | 0.779 | 0.660 | 0.642 | 0.601 | 0.516 |
| **kappa score** | 0.319 | 0.378 | 0.299 | 0.304 | 0.257 | 0.218 |

**Figure 6.**
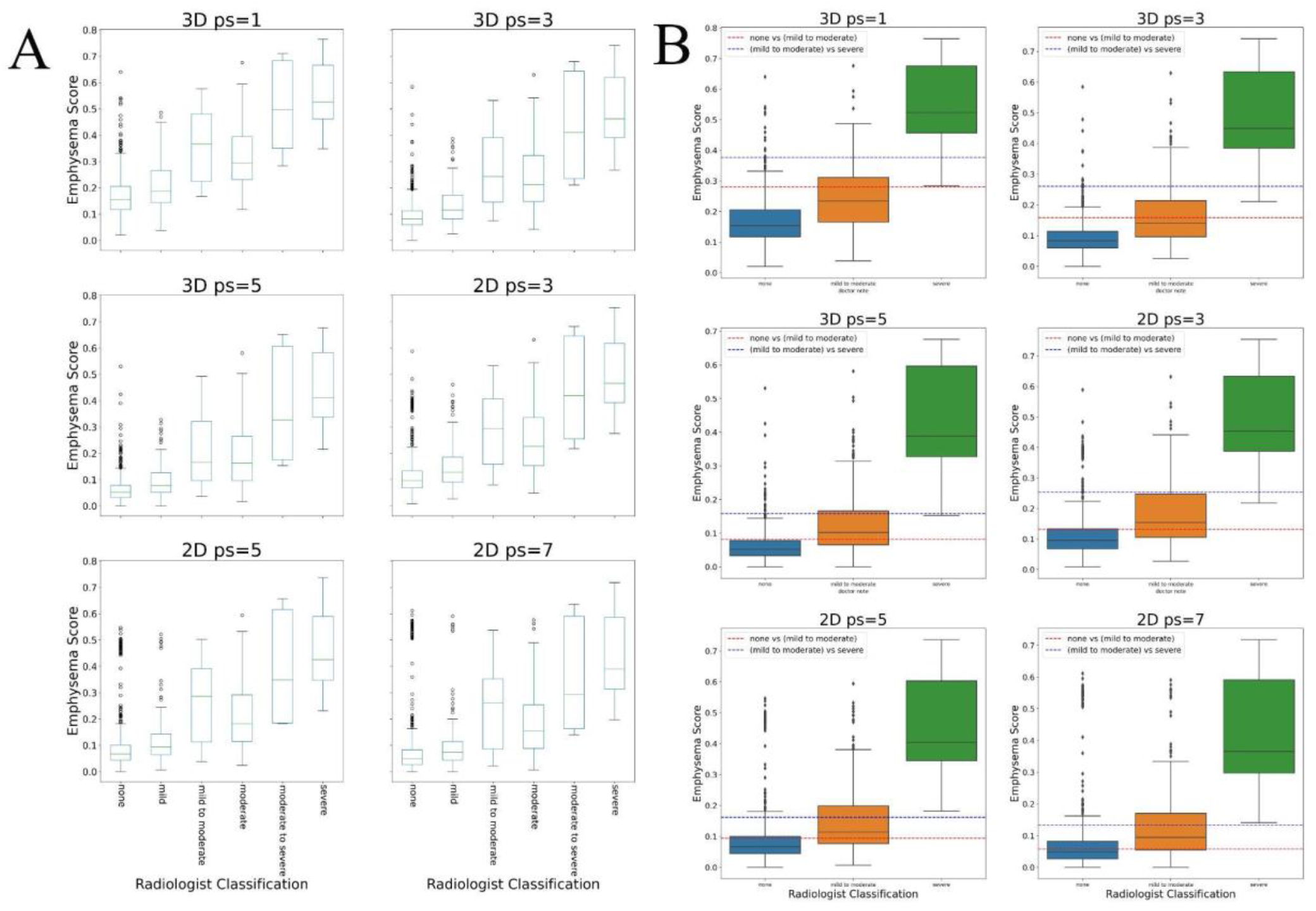
(A) Cubic root transformed emphysema score distributions of the external validation dataset of 6 different kernels. (B) Cubic root transformed emphysema score distributions after regrouping with the final cutoffs.

## 4. Discussion

We have demonstrated an accurate emphysema quantification algorithm that leverages 3-dimensional multi-voxel kernels and image processing with airways removal. The 3×3×3 (3D *ps*=3) kernel (AUC = 0.782 & 0.985, accuracy=0.676 & 0.757) achieved accurate emphysema quantification as demonstrated by the provided evaluation metrics and qualitative analyses of heat map overlays of emphysema regions.

Accurate emphysema quantitation has a number of important clinical and research applications. First, the current practice of reporting emphysema in clinical radiology is subjective and poorly calibrated among radiologists at different practices. Quantitative imaging would introduce more reliable reporting of emphysema and its severity in radiology reporting. Furthermore, being able to objectively quantify emphysema opens up the possibility of more precisely defining a research cohort (e.g. severe emphysema cohort as quantified by imaging) rather than relying on subjective classification by a radiologist. Finally, being able to identify an area with objective severe emphysema would allow for more precise targeting and treatment of obstructive airways disease by lung reduction surgery.

Traditionally accepted emphysema quantitation by counting voxel-wise LAA had a number of disadvantages including overcounting of quantum mottles in low dose CT as well as subclinical emphysema that likely contribute minimally to the overall disease severity. By counting in 2D or 3D kernels in patches, our proposed algorithm mitigates these shortcomings and focuses on larger and connected regions of emphysema.

In evaluating our algorithm, the 2D kernels achieved comparable AUC, accuracy, and other metrics with 3D *ps*=3 kernel, but the score distribution was less robust in the external validation dataset, resulting in lower performance than the LAA score. Indeed, in cases where one particular axial slice is especially noisy or the lung segmentation has failed to remove airways very close to lung lobes, 3D kernels ignored this false positive if its neighboring axial slices are normal, but such conditions inflated the emphysema scores produced by 2D kernels, which neglected the connection along the inferior-superior axis. Although the 3D *ps*=1 (LAA score) also achieved comparable accuracy with the 3D *ps*=3 kernel on the external validation, the former’s low AUC scores make it unlikely to be a good predictor. Furthermore, inspecting the confusion matrix and the boxplot for the external validation dataset showed that the LAA score was more biased toward predicting “none” classes than the 3D *ps*=3 kernel.

A few notable incorrect predictions from error analysis included false positive cases resulting from pulmonary cysts, honeycombing, and bronchiolitis obliterans. The algorithm not being able to differentiate false-positive pathologies in CT scans from true emphysema is not unexpected given that these pathologies also present with areas of low attenuation. These may ultimately need to be resolved at the discretion of clinical radiologists or with more advanced deep learning based algorithms that may be able to take into account contextual information surrounding the pathologies.

A few limitations are noted for the study. First, the ground truth labels of emphysema severity extracted from radiology reports were sometimes imperfect, especially in the cases where there was no mention of emphysema. Most cases that did not mention emphysema were noted to have little to no emphysema based on our quantitation algorithm, but the cost of re-annotating the images was prohibitive for the study. Second, the ground truth labels were based on radiologic classification of emphysema rather than clinical COPD severity assessment tools such as the GOLD criteria. However, GOLD criteria may be also influenced by chronic bronchitis in addition to the degree of emphysema, therefore, arguably, no existing clinical guideline would be perfect in quantifying the degree of lung parenchymal destruction. Third, since the lung segmentation component of this algorithm removes small patches of dense regions within the lung lobe that are below −800 HU during the airway removal process, it occasionally over-removes small lung parenchyma regions that are emphysema-rich and connected to major airways. However, given that these cases represent severe emphysema patients, such slight over-removal of emphysematous regions were unlikely to affect the final emphysema severity classification.

In conclusion, we propose a 3D averaging kernel based algorithm that can quantify and differentiate emphysema severity, which can be useful for integrating quantitative imaging and its associated objectivity into radiology reporting, which in turn can better inform lung cancer risk, define research cohorts, and guide lung reduction surgery planning.

## Supporting information

Supplemental formulae, figures, and tables mentioned in the manuscript.

STARD checklist

## Data Availability

Data produced by this study contains confidential information about patients and is not publicly available. Codes accompanying this study are available at https://github.com/bdrad/Emphysema_Quantification

## Author Contributions

Conceptualization, JZ, JHS; methodology, JZ, JHS, GRC, MB; validation, JZ, JHS, GRC, MB; resources, JHS; data curation, JZ, JHS, GRC; writing, JZ, JHS, GRC, MB; code and visualization, JZ.

