## Supplemental formulae, figures, and tables mentioned in the manuscript. for "Emphysema Quantification and Severity Classification with 3-Dimensional Averaging Kernel and Airways Removal"

### Supplementary Information

#### *Mathematical Derivations*

For each voxel coordinate $(z, x, y)$, the Hounsfield unit at this coordinate is represented by the mapping $H(z, x, y)$. Define M as the set of voxel coordinates in the lung segmentation mask. Consider an arbitrary $(z_0, x_0, y_0)$, let S denote the set of voxel coordinates within the patch centered by $(z_0, x_0, y_0)$ with patch size p (only odd patch sizes are considered here) is defined as: $S_{3D}(z_0, x_0, y_0) = \{(z, x, y) \mid |z - z_0| \leq \frac{p-1}{2} \wedge |x - x_0| \leq \frac{p-1}{2} \wedge |y - y_0| \leq \frac{p-1}{2}\}$ in the 3D case, and $S_{2D}(z_0, x_0, y_0) = \{(z, x, y) \mid z = z_0 \wedge |x - x_0| \leq \frac{p-1}{2} \wedge |y - y_0| \leq \frac{p-1}{2}\}$ in the 2D case.

The mapping from $(z_0, x_0, y_0)$ to its value in the patch average is defined as:

$P(z_0, x_0, y_0) = \frac{\sum_{(z,x,y) \in S(z_0, x_0, y_0)} H(z,x,y) I((z,x,y) \in M)}{\sum_{(z,x,y) \in S(z_0, x_0, y_0)} I((z,x,y) \in M)}$ where *I* is the indicator function, which allows only voxels within the lung mask to contribute to the mapped value so that the high Hounsfield value of the tissue near the periphery of the lung will not affect emphysema quantification. The final emphysema score is defined as:

$ES = \frac{\sum_{(z,x,y) \in M} I(P(z,x,y) < -950)}{\sum_{(z,x,y) \in M} 1}$ that represents the fraction of voxels in the lung mask that is below -950 HU.

### *Figures and Tables*

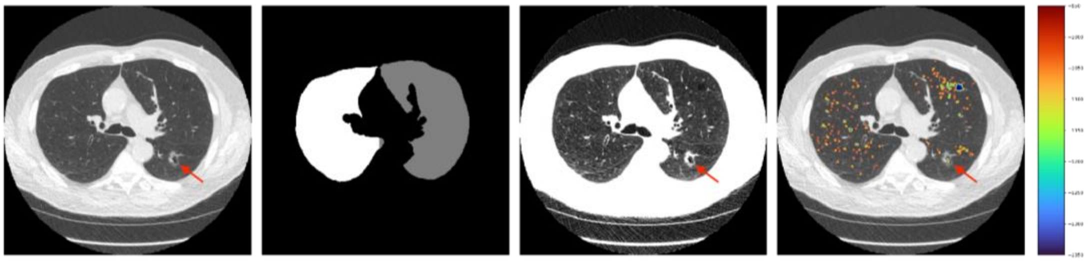

**Figure S1. Emphysema quantification false positive case.** Pulmonary cyst shown by the red arrow was included in the emphysema mask. Column 1-4: axial ct scan with window level -600 and window width 1500, lung mask, axial ct scan with window level -750 and window width 600, axial ct scan overlayed with heatmap indicating emphysema locations and severity as classified by 3D kernel with patch size = 3 (i.e. a 3x3x3 kernel).

| | 3D ps=1 | | | 3D ps=3 | | | 3D ps=5 | | |
|---|---|---|---|---|---|---|---|---|---|
| | none | mild to moderate | severe | none | mild to moderate | severe | none | mild to moderate | severe |
| none | 338.8 | 86.1 | 47.1 | 387.7 | 61.6 | 22.7 | 381.9 | 73.9 | 16.2 |
| mild to moderate | 115.4 | 60.9 | 48.7 | 111.3 | 69.2 | 44.5 | 123.4 | 58.4 | 43.2 |
| severe | 0 | 3.5 | 21.5 | 0 | 0.4 | 24.6 | 0.6 | 0.4 | 24 |

**Table S1.** Average Confusion Matrix for Cutoff Determination Dataset

| | 3D ps=1 | | | 3D ps=3 | | | 3D ps=5 | | |
|---|---|---|---|---|---|---|---|---|---|
| | none | mild to moderate | severe | none | mild to moderate | severe | none | mild to moderate | severe |
| none | 674 | 48 | 14 | 669 | 52 | 15 | 564 | 144 | 28 |
| mild to moderate | 155 | 45 | 35 | 129 | 60 | 46 | 94 | 80 | 61 |
| severe | 0 | 3 | 32 | 0 | 2 | 33 | 0 | 1 | 34 |

**Table S2.** Confusion Matrix for External Validation Dataset.
