## Supplementary material for "Emphysema Quantification and Severity Classification with 3-Dimensional Averaging Kernel and Airways Removal": STARD checklist

### STARD 2015 Checklist

| Section & Topic | No. | Item | ✔ (N/A) |
|---|---|---|---|
| **TITLE OR ABSTRACT** | | | |
| | 1 | Identification as a study of diagnostic accuracy using at least one measure of accuracy (such as sensitivity, specificity, predictive values, or AUC) | ✔ |
| **ABSTRACT** | | | |
| | 2 | Structured summary of study design, methods, results, and conclusions (for specific guidance, see STARD for Abstracts) | ✔ |
| **INTRODUCTION** | | | |
| | 3 | Scientific & clinical background, including the intended use and clinical role of the index test | ✔ |
| | 4 | Study objectives and hypotheses | ✔ |
| **METHODS** | | | |
| ***Study design*** | 5 | Whether data collection was planned before the index test and reference standard were performed (prospective study) or after (retrospective study) | ✔ |
| ***Participants*** | 6 | Eligibility criteria | ✔ |
| | 7 | On what basis potentially eligible participants were identified (such as symptoms, results from previous tests, inclusion in registry) | ✔ |
| | 8 | Where and when potentially eligible participants were identified (setting, location and dates) | ✔ |
| | 9 | Whether participants formed a consecutive, random or convenience series | ✔ |
| ***Test methods*** | 10a | Index test, in sufficient detail to allow replication | ✔ |
| | 10b | Reference standard, in sufficient detail to allow replication | ✔ |
| | 11 | Rationale for choosing the reference standard (if alternatives exist) | ✔ |
| | 12a | Definition of and rationale for test positivity cut-offs or result categories of the index test, distinguishing pre-specified from exploratory | ✔ |
| | 12b | Definition of and rationale for test positivity cut-offs or result categories of the reference standard, distinguishing pre-specified from exploratory | ✔ |
| | 13a | Whether clinical information and reference standard results were available to the performers/readers of the index test | ✔ |
| | 13b | Whether clinical information and index test results were available to the assessors of the reference standard | ✔ |
| ***Analysis*** | 14 | Methods for estimating or comparing measures of diagnostic accuracy | ✔ |
| | 15 | How indeterminate index test or reference standard results were handled | ✔ |
| | 16 | How missing data on the index test and reference standard were handled | ✔ |
| | 17 | Any analyses of variability in diagnostic accuracy, distinguishing pre-specified from exploratory | ✔ |
| | 18 | Intended sample size and how it was determined | N/A |
| **RESULTS** | | | |
| ***Participants*** | 19 | Flow of participants, using a diagram. Include the figure number (preferably figure 1) or page number | ✔ |
| | 20 | Baseline demographic and clinical characteristics of participants | ✔ |
| | 21a | Distribution of severity of disease in those with the target condition | ✔ |
| | 21b | Distribution of alternative diagnoses in those without the target condition | N/A |
| | 22 | Time interval and any clinical interventions between index test and reference standard | N/A |
| ***Test results*** | 23 | Cross tabulation of the index test results (or their distribution) by the results of the reference standard | ✔ |
| | 24 | Estimates of diagnostic accuracy and their precision (such as 95% confidence intervals) | ✔ |
| | 25 | Any adverse events from performing the index test or the reference standard | N/A |
| **DISCUSSION** | | | |
| | 26 | Study limitations, including sources of potential bias, statistical uncertainty, and generalisability | ✔ |
| | 27 | Implications for practice, including the intended use and clinical role of the index test | ✔ |
| **OTHER INFORMATION** | | | |
| | 28 | Registration number and name of registry | N/A |
| | 29 | Where the full study protocol can be accessed | N/A |
| | 30 | Sources of funding and other support; role of funders | ✔ |

*N/A stands for not applicable and may be a reasonable choice depending on the type of study performed
